# Variant-specific antibody correlates of protection against SARS-CoV-2 Omicron symptomatic and overall infections

**DOI:** 10.1101/2025.02.11.25322066

**Authors:** Jose Victor Zambrana, Ian A. Mellis, Abigail Shotwell, Hannah E. Maier, Yara Saborio, Carlos Barillas, Roger Lopez, Gerald Vasquez, Miguel Plazaola, Nery Sanchez, Sergio Ojeda, Isabel Gilbertson, Guillermina Kuan, Qian Wang, Lihong Liu, Angel Balmaseda, David D. Ho, Aubree Gordon

**Affiliations:** Department of Epidemiology, University of Michigan, Ann Arbor, MI; Sustainable Sciences Institute, Managua, Nicaragua; Aaron Diamond AIDS Research Center, Vagelos College of Physicians and Surgeons, Columbia University Irving Medical Center, New York, NY; Department of Pathology and Cell Biology, Vagelos College of Physicians and Surgeons, Columbia University Irving Medical Center, New York, NY; New York Blood Center, New York, NY; Laboratorio Nacional de Virología, Centro Nacional de Diagnóstico y Referencia, Ministerio de Salud, Managua, Nicaragua; Centro de Salud Sócrates Flores Vivas, Ministerio de Salud, Managua, Nicaragua

**Author notes:** Correspondence: Aubree Gordon. Department of Epidemiology, University of Michigan, Ann Arbor, 5622 SPH I 1415 Washington Heights Ann Arbor, MI 48109., Alternate Correspondence: David D. Ho, Aaron Diamond AIDS Research Center, Columbia University Irving Medical Center, 701 West 168th Street, HHSC 1102, New York, NY 10032. Jose Victor Zambrana and Ian A. Mellis contributed equally to this manuscript.

## Abstract

**Background:** Vaccination and prior infection elicit neutralizing antibodies targeting SARS-CoV-2, yet the quantitative relationship between serum antibodies and infection risk against viral variants remains uncertain, particularly in underrepresented regions.

**Methods:** We investigated the protective correlation of pre-exposure serum neutralizing antibody levels, employing a panel of SARS-CoV-2 pseudoviruses (Omicron BA.1, Omicron BA.2, and ancestral D614G), and Spike-binding antibody levels, with symptomatic BA.1 or BA.2 SARS-CoV-2 infections and overall infection, in 345 household contacts from a SARS-CoV-2 household cohort study.

**Results:** A four-fold increase in homotypic-neutralizing (e.g., BA.1-neutralizing vs. BA.1 exposure) titers was correlated with protection from symptomatic infections (BA.1 protection: 28% [95%CI 12–42%]; BA.2 protection: 43% [20–62%]), and ancestral-neutralizing titers were also correlated with protection from either variant, but only at higher average levels than homotypic. Mediation analyses revealed that homotypic and D614G-neutralizing antibodies mediated protection from infection and symptomatic infection both from prior infection and vaccination.

**Conclusions:** These findings underscore the importance of monitoring variant-specific antibody responses and highlight that antibodies targeting circulating strains may be more predictive of protection from infection. Nevertheless, ancestral-strain-neutralizing antibodies remain relevant as a correlate of protection. Our study emphasizes the need for continued efforts to assess antibody correlates of protection.

**Funding:** We acknowledge funding from the U.S. N.I.H., the Open Philanthropy Project, and the Bill and Melinda Gates Foundation.

**Research In Context:** 

**Evidence before this study:** Based on searches of Google Scholar and PubMed, not restricted to English-language articles, using search terms including “correlates of protection,” “SARS-CoV-2,” “COVID-19,” “BA.1,” “BA.2,” “neutralizing antibodies,” “immune response,” and “thresholds of protection” we identified multiple studies, primarily based on randomized clinical trials or prospective cohort designs, which have shown that serum SARS-CoV-2-neutralizing antibodies are informative correlates of protection from overall or symptomatic infection after vaccination or infection. Our search was limited to 2020 and later, and included several high-quality RCTs and cohort studies, as well as consideration of published meta-analyses. Often the serum correlates used were spike-binding or -neutralizing peak titers or baseline titers months before infection waves, and specific viral exposures for participants remained unknown. Most prior evidence related to neutralizing antibodies focused on neutralizing titers against the ancestral strain even when participants were challenged with later viral variants, although a small number of groups have also investigated neutralizing antibody titers directed against contemporaneous strains, such as Omicron BA.1. Furthermore, most studies of correlates of protection focused on vaccines widely available in North America and Europe; few serum antibody correlates of protection studies have included globally used COVID-19 vaccines, such as Soberana or Sputnik. Relatedly, Central America as a region has had no known studies of SARS-CoV-2 serum neutralizing antibody correlates of protection, to the best of our knowledge, despite the unique vaccine exposure histories of its residents and widespread infections before such vaccines became available.

**Added value of this study:** In this study, we used a household-based cohort study design with an embedded transmission study, in which we collected serum from household members just before they were exposed to a co-resident in the household who developed COVID-19. This design enabled correlates of protection analysis of serum antibody titers at the time of known exposure, relatively homogeneously across the cohort. Our study was conducted in Managua, Nicaragua, during Omicron BA.1 and BA.2 infection waves, presenting the first such study in Central America, and the first such study including participants who received some global vaccines. We conducted pseudovirus neutralization assays against the ancestral SARS-CoV-2 strain and contemporaneous Omicron BA.1 and BA.2 variants, allowing for parallel analyses of anti-Omicron titers as correlates of protection and more widely available ancestral-neutralizing titers.

**Implications of all the available evidence:** Overall, the results in this study, in combination with the prior available evidence, continue to point to serum neutralizing antibodies as an informative correlate of protection from overall and symptomatic infection. Titers against contemporaneous variants (BA.1 and BA.2 in this case), were protective at lower levels than those against the ancestral strain, but ancestral-neutralizing titer was still informative as a correlate of protection. Lastly, for any known vaccination or prior infection history studied with such an analysis, neutralizing antibodies appear to mediate protective effects of prior SARS-CoV-2 exposure on overall or symptomatic infection.

## Background

Some people exposed to SARS-CoV-2 become infected shortly thereafter, while others do not. Humoral immunity, elicited by prior infection or vaccination, is important for protection from infection.^1^ At a population level, several groups have shown that serum anti-SARS-CoV-2 antibody titers are an informative correlate of protection from infection in clinical trials and observational cohorts.^2-9^ However, the extent to which serum anti-SARS-CoV-2 antibody titers correlate with protection is highly variable, explaining 48.5% to 94.2% of infection risk across different vaccine efficacy studies.^10^ Additionally, SARS-CoV-2 continues to evolve and increasingly evade existing antibody responses.^11-13^ The extent to which such evolution compromises the interpretation of anti-SARS-CoV-2 antibody assays focused on historical, non-predominant variants remains underexplored.^14^ Also of importance to global public health, the majority of published studies of antibody correlates of protection focus on participants from North America, Europe, and Asia; relatively few include participants from Central and South America or Africa where a majority of first SARS-CoV-2 exposures were through infection, not vaccination, and less-studied vaccines were deployed.^15^

In most studies of correlates of protection from infection, specific participant virus exposures remain unidentified, and individuals will vary widely in their exposures over time, unless prevalence is very high. Herein, we overcome this tracking problem with a household-based cohort study with an embedded transmission study, intensely monitoring “index cases” who present with infection and their household member “contacts,” building on our prior study, in which we identified only clinical correlates of protection from SARS-CoV-2 infection in households in Nicaragua.^16^ The present study aims to measure protection against SARS-CoV-2 BA.1 and BA.2 infection and moderate/severe infection in the Household Influenza Cohort Study (HICS) population following transmission of SARS-CoV-2 (prior infections with ancestral or Delta strains) and vaccinations. We combine household-level infection tracking data with hundreds of participants’ neutralizing antibody titers against ancestral SARS-CoV-2 (D614G) and the contemporaneous exposure viral variants Omicron BA.1 and BA.2, spike-binding titers, and clinical information, including infection and vaccination histories and age, to better understand clinical and serum antibody measurements that are predictive of protection from infection or symptomatic infection risk after a given virus exposure. Finally, we integrate infection and vaccination histories with neutralizing titers in causal mediation analysis to show that neutralizing antibodies mediate protective effects of prior viral exposures from infection or vaccination.

## Methods

### Study design and participants

The Household Influenza Cohort Study (HICS) is an ongoing prospective cohort study in District II of Managua, Nicaragua in the Health Center Socrates Flores Vivas (HCSFV). Established in 2017 to investigate influenza, it was expanded in early 2020 to include SARS-CoV-2 infection and disease.^16^ All cohort participants undergo regular blood sampling—twice per year in 2020 and 2021, and then annually thereafter. At the first sign of any illness, participants are instructed to seek care at the study health clinic, where they receive primary care and diagnostic testing (Figure S1).

An embedded household transmission study is activated once a participating household member is confirmed positive for SARS-CoV-2 (Figure S1). After activation, study personnel visit the household on days 0, 3, 7, 14, 21, and 30 to collect combined nasal/oropharyngeal swabs, maintain symptom diaries, and test all household members regardless of symptoms. Serum samples are collected on the same day as activation. Active surveillance of household contacts allows the detection of symptomatic and asymptomatic SARS-CoV-2 infections.

These studies received approval from the institutional review boards of the Nicaraguan Ministry of Health and the University of Michigan (HUM00119145 and HUM00178355). Written informed consent or parental permission was obtained for all enrolled participants, and children aged 6 years or older also provided assent.

### Inclusion and exclusion criteria

Households were eligible for inclusion if they experienced an introduction of either BA.1 or BA.2 SARS-CoV-2 strains, which circulated sequentially from January and June 2022 (Figure S2). Pre-exposure samples were included in the analysis if they were collected from 10 days before up to 4 days after the household’s activation date. If a participant lacked a sample within this window but had one collected within the proceeding 90 days-without documented SARS-CoV-2 infection or vaccination in the interim- and before the first positive respiratory sample in the activation period, it was also accepted as a pre-exposure sample. Based on these criteria, 14 participants were excluded.

Strain ascertainment was performed by direct sequencing from household members or by single imputation using weekly sequencing data from the Ministry of Health of Nicaragua (Managua department) (Figure S4). Two households (totaling 11 individuals) were subsequently excluded because they lacked sufficient data for direct sequencing or single imputation (multiple strains were circulating that week). In total, these criteria resulted in a final cohort of 345 participants, 251 within the BA.1 Omicron wave and 94 individuals within the BA.2 Omicron wave.

### Laboratory methods

Blood samples were analyzed in pairs (current vs. baseline) using enzyme-linked immunosorbent assay (ELISA) protocols adapted from the Krammer laboratory for the detection of antibodies against SARS-CoV-2 spike receptor binding domain (RBD), Spike and Nucleoprotein (NP). The RBD and spike proteins used in these assays were produced in single batches at the Life Sciences Institute at the University of Michigan, based on the original SARS-CoV-2 strain. Due to its specificity, RBD was used for initial screening (positive/negative), while positive samples were subsequently titrated using the spike assay. The NP ELISA was also performed to differentiate vaccine-induced responses from those due to infection. Real-time reverse-transcription polymerase chain reaction (RT-PCR) was conducted following the protocol described by Chu et al.^17^

Neutralization titers against BA.1, BA.2 and D614G SARS-CoV-2 strains were measured using vesicular stomatitis virus–based pseudoviruses (VSV) bearing different SARS-CoV-2 spike proteins produced in HEK293T cells, as previously described.^11^ After transfecting spike-encoding plasmids, a VSV-G pseudotyped ΔG-luciferase was used to generate the pseudoviruses, which were harvested and standardized by determining the 50% tissue culture infectious dose (TCID50) in Vero-E6 cells. Heat-inactivated serum samples were then serially diluted, incubated with each pseudovirus, and added to Vero-E6 cells, followed by a 16-hour incubation. Luciferase activity was subsequently measured, and ID50 values for each strain-specific pseudovirus were calculated using a five-parameter log-logistic model to compare neutralization responses across the different SARS-CoV-2 variants. Additional details are included in the supplementary material.

### Endpoints

Symptoms reported in daily symptom diaries and during clinic visits were used to categorize illness severity into four levels. “Severe” cases included individuals requiring hospitalization, and those deemed in need of hospitalization by clinicians. No deaths due to COVID-19 were recorded during the Omicron BA.1 or BA.2 waves in the HICS study. “Moderate” cases encompassed individuals presenting with difficulty breathing, shortness of breath, rapid breathing, tight chest feeling, chest pain, ALRI, SARI, crepitus, chest wall indrawing, rhonchi, wheezing, or an overall poor condition. “Mild” cases were defined as those with loss of smell/taste, fever, or at least two other symptoms. Finally, “subclinical” cases involved no more than one symptom that did not meet the criteria for higher severity. The primary endpoints of this study were the percentage protection against RT-PCR-confirmed infection—including subclinical, mild, moderate, and severe infections, (hereafter referred to as ‘infection’)—and symptomatic infection (including mild, moderate, and severe infections) among households with SARS-CoV-2 BA.1 or BA.2 exposure.

### Independent variables

Antibody titers (the primary independent variables of interest) across assays were log-transformed using base 4 (log4[titer]). Assays included D614G, BA.1 and BA.2 neutralization tests, and Spike ELISA. Infectious dose 50s (ID50s) were calculated using a 5-parameter log-logistic model among the neutralization assays. A modification of the Reed and Muench formula was used to calculate titers for the Spike ELISA.^18^ Confounders included age (continuous variable), vaccination status (any prior vaccination, dichotomous variable) and infection history (any prior infection, dichotomous variable). Infection history and vaccination ascertainment details are included as supplementary materials. The selection of confounders was based on directed acyclic graph (DAG) analysis supported by the existing literature (Figure S3).

### Statistical analysis

Univariate analyses using Wilcoxon rank sum test were applied to examine the relationship between antibody titer and infection outcome by assay and Omicron wave (BA.1 or BA.2). We also assessed the multivariate association between titers and infection outcomes by Omicron wave and assay with generalized linear models (GLMs) under a binomial family. Protection associated with increasing titer levels was quantified as 1 – odds ratio (OR). We also determined protective cut-offs at 50 and 80% using GLM and Generalized Additive Models (GAM) with thin-plate spline regression with three knots to allow for non-linear effects.

Multivariate GAM and GLM models were adjusted for the confounders mentioned above.

We also conducted a causal mediation analysis, as a secondary analysis, to examine how neutralizing antibodies mediate protection from prior vaccination and natural infection against both overall and symptomatic infection, using a GLM framework with non-parametric bootstrapping and accelerated bias-corrected 95% confidence intervals applying 1000 simulations.

All analyses examining the likelihood of infection or symptomatic infections included the computation of 95% confidence intervals (CIs) using standard errors, assuming a normal distribution with an α level of 0.05, except for the mediation analyses. Statistical significance was evaluated using two-tailed Wald tests on the regression coefficients. All statistical analyses were conducted using R (R Foundation for Statistical Computing v4.3.1) and mediation statistical package (v4.5.0) (cite).^19,20^

## Results

We measured SARS-CoV-2 antibody levels in 345 individuals from the HICS study before exposure to BA.1 or BA.2 and used these measurements to assess the protective effect of pre-existing antibodies against infection and moderate/severe infection. Households included in the study experienced the introduction of BA.1 or BA.2 SARS-CoV-2 strains by a household member, with these strains circulating consecutively between January and June 2022 (Figure S2). The final sample consisted of 194 individuals who were later infected by either the BA.1 (N = 153) or BA.2 (N = 41) strains, and 151 individuals who remained uninfected. Study participants had a median age of 18 years (IQR 9–40), with a majority being female (211/345 [61%]). By the time of sampling, most participants experienced at least one prior SARS-CoV-2 infection (322/345 [93%]) and had received at least one vaccine dose (233/345 [68%]) (Table 1). The average number of prior vaccine doses among individuals sampled before BA.2 introduction compared to those sampled before BA.1.

**Table 1.** Participant characteristics.

| Characteristic | Overall | BA.1 wave | BA.2 wave |
| --- | --- | --- | --- |
| Participants – N | 345 | 251 | 94 |
| Households – N | 91* | 66 | 32 |
| Participants per household – Median (IQR) | 3 (2, 5) | 3 (2, 5) | 3 (2, 3) |
| Age – Median (IQR) | 18 (9, 40) | 18 (9, 39) | 17 (10, 41) |
| Female sex – N (%) | 211 (61%) | 151 (60%) | 60 (64%) |
| Infected – N (%) | 194 (56%) | 153 (61%) | 41 (44%) |
| Infection severity – N (%) |  |  |  |
| Subclinical | 89 (46%) | 77 (50%) | 12 (29%) |
| Mild | 78 (40%) | 52 (34%) | 26 (63%) |
| Moderate | 24 (12%) | 21 (14%) | 3 (7.3%) |
| Severe | 3 (1.5%) | 3 (2.0%) | 0 (0%) |
| Infection history – N (%) |  |  |  |
| No prior infections | 23 (6.7%) | 18 (7.2%) | 5 (5.3%) |
| 1 prior infection | 235 (68%) | 171 (68%) | 64 (68%) |
| 2 prior infections | 80 (23%) | 58 (23%) | 22 (23%) |
| 3 prior infections | 7 (2.0%) | 4 (1.6%) | 3 (3.2%) |
| Time since last infection – Median (IQR) | 406 (163, 584) | 380 (150, 580) | 437 (263, 689) |
| Vaccination history |  |  |  |
| Unvaccinated | 112 (32%) | 90 (36%) | 22 (23%) |
| 1 prior dose | 71 (21%) | 59 (24%) | 12 (13%) |
| 2 prior doses | 75 (22%) | 57 (23%) | 18 (19%) |
| >2 prior doses | 87 (25%) | 45 (18%) | 42 (45%) |
| Time since last Vax dose – Median (IQR) | 77 (34, 102) | 62 (22, 93) | 112 (77, 162) |
| Pre-infection titers – Median (IQR) |  |  |  |
| Neut BA.1 ID<sub>50</sub> | 1347 (180, 3330) | 1061 (146, 3130) | 1839 (554, 3943) |
| Neut BA.2 ID<sub>50</sub> | 1482 (292, 4517) | 1332 (218, 3975) | 2150 (709, 5338) |
| Neut DG14G ID<sub>50</sub> | 5038 (1315, 11694) | 4798 (1173, 12509) | 5132 (1954, 10882) |
| Spike ELISA titer | 8553 (2198, 28937) | 7166 (1505, 26138)** | 10660 (5168, 36256) |
| Sample time before infection (Neut)*** – Median (IQR) | -4 (-17, 0) | -4 (-14, 0) | -3 (-22, 0) |
| Sample time before infection (Spike)*** – Median (IQR) | -6 (-60, 0) | -6 (-67, 0)** | -6 (-60, 0) |

\*91 unique households. 7 households were activated in both waves.
\*\*1 individual with unknown titer
\*\*\*Among BA.1 or BA.2 infected participants; "Neut": samples for neutralizing titers; "Spike": for spike-binding.

We measured serum neutralization of BA.1, BA.2, and D614G SARS-CoV-2 using pseudovirus assays, and assessed Spike-binding titers using ELISA. Samples were collected prior to infection or up to 4 days after the first RT-PCR-positive detection within a household with a median of -4 days (Interquartile range [IQR] -17–0) relative to the first positive detection date for the neutralization assays, and -6 days (IQR -60–0) for Spike differing by sample availability and with 84% samples shared across assays (Table 1). We found that all measured immune markers were consistently higher before the BA.2 introduction compared to the BA.1 introduction (Table 1). While all markers measured in the study were co-linear at the time of sampling, the greatest collinearity was observed between BA.1- and BA.2-specific titers (Figure S4). We then evaluated how these markers correlate with outcomes. In our univariate analyses, neutralizing titers across viruses were significantly associated with protection from infection with BA.2 but not BA.1 (Table S1, Figure S5). In contrast, neutralizing titers were significantly associated with protection from symptomatic infections with BA.1 and BA.2 infections, except for D614G titers in relation to symptomatic BA.1 infection (P-value 0.106) (Table S2, Figure S6).

Similarly, after adjusting for age, infection history and vaccine history, we found that a four-fold linear increase in neutralizing titers across neutralization assays was significantly associated with protection from BA.2 infection—45% (95% Confidence Interval [CI] 21–64%) for BA.1 neutralizing titers, 45% (CI 21–64%) for BA.2 neutralizing titers, and 51% (CI 23–71%) for D614G neutralizing titers—but not with protection from BA.1 infection (Figure S7, Table S3). Spike-binding titers were not associated with protection from BA.1 or BA.2 infection (Figure S7, Table S3). However, both neutralizing and spike-binding titers were associated with protection from symptomatic infection across waves (Figure S7, Table S4). Although similar levels of protection were observed for each four-fold increase in neutralizing titers across assay, the average pre-exposure titer values were higher for D614G than for the Omicron sub variants (Figure S7, Table 1).

We then analyzed infections and symptomatic infections in the range of observable titers among vaccinated and previously infected adults to derive protection thresholds for BA.1 and BA.2 infection outcomes. Titers to achieve 50% and 80% protection from BA.2 infections were similar for BA.1 and BA.2 neutralizing titers. However, achieving the same level of protection using D614G neutralizing titers required more than three times higher than BA.1 or BA.2 neutralizing titers (Figure 1A, Table S5-6). In contrast, titers required for 50% or 80% protection from symptomatic BA.1 or BA.2 infection were lower than those needed to prevent BA.2 infection. Similarly, protective titers against contemporaneous strains were lower compared to D614G neutralizing titers (Figure 1B, Table S5-6). For instance, achieving 50% protection from BA.1 infection required a BA.1 neutralizing titer of 39 and BA.2 neutralizing of 53, whereas a D614G neutralizing titer of 222 was needed. Protective titers based on Spike-binding assays were also reported (Figure S8).

**Figure 1.**
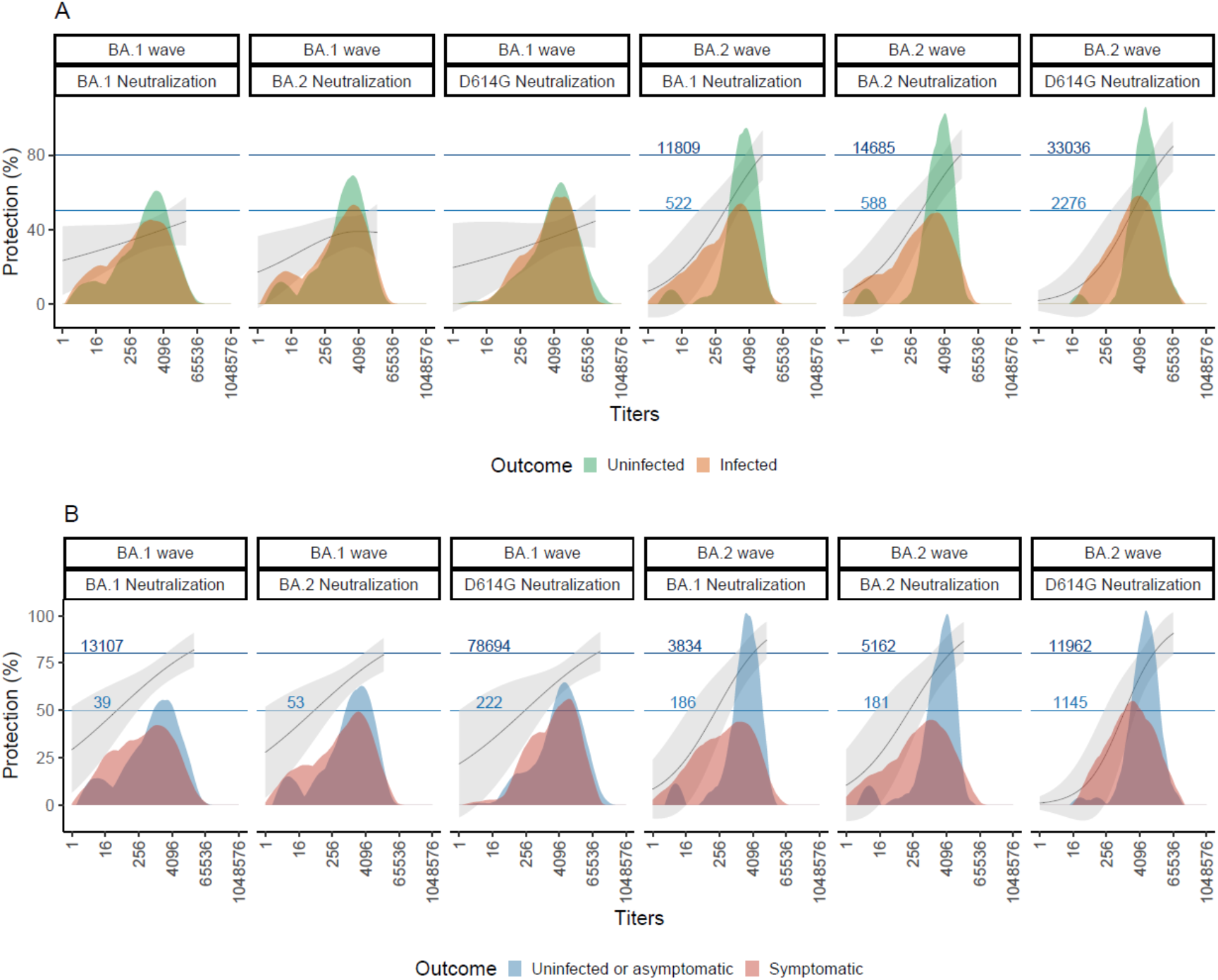
Neutralizing titers as a correlate of protection by Omicron wave and assay. Relationship between neutralizing titers (ID_50_) and protection against BA.1 and BA.2 infection (**A**) and symptomatic infection (**B**) during their respective waves. The solid lines represent the predicted protection curves according to a Generalized Additive Model (see Methods), with the shaded ribbon bands denoting the 95% confidence intervals (CIs) for these estimates, derived from vaccinated and prior-infected adults. The horizontal blue solid lines mark the points where the predicted protection curves cross the 50% and 80% protection thresholds, highlighting the titer levels required to achieve these levels of protection. Embedded within the figure are histograms showing the raw distribution of neutralization titers, stratified by outcome.

We next employed causal mediation analysis to evaluate the role of antibody responses in mediating the protective effects of prior infection or vaccination against subsequent infection and symptomatic infection. For these analyses, we combined both Omicron waves and adjusted for Omicron wave, age and prior infection or vaccination (as main exposure or a confounder, as appropriate). “Homotypic” antibodies (e.g., BA.1- or BA.2-neutralizing antibodies against BA.1 or BA.2 infections, respectively) significantly mediated protection, with mediation effect estimates indicating that every four-fold increase in titer mediated 11% (CI 4–21%, p = 0.006) and 12% (CI 6–24%, p = 0.002) protection from prior infection for infection and symptomatic infection, respectively (Figure 2, Table S7). Additionally, for every four-fold increase in titer, “homotypic” neutralizing antibodies significantly mediated 5% (CI 2–9%, p < 0.001) protection against infection and 7% (CI 4–11%, p < 0.001) protection against symptomatic infection from vaccination. D614G neutralizing antibodies also significantly mediated protection from prior infection and vaccination, though with slightly lower mediation effects. In contrast, D614G spike-binding antibodies did not significantly mediate protection against infection and only mediated significant protection against symptomatic infection from both vaccination and prior infection (Figure 2, Table S7).

**Figure 2.**
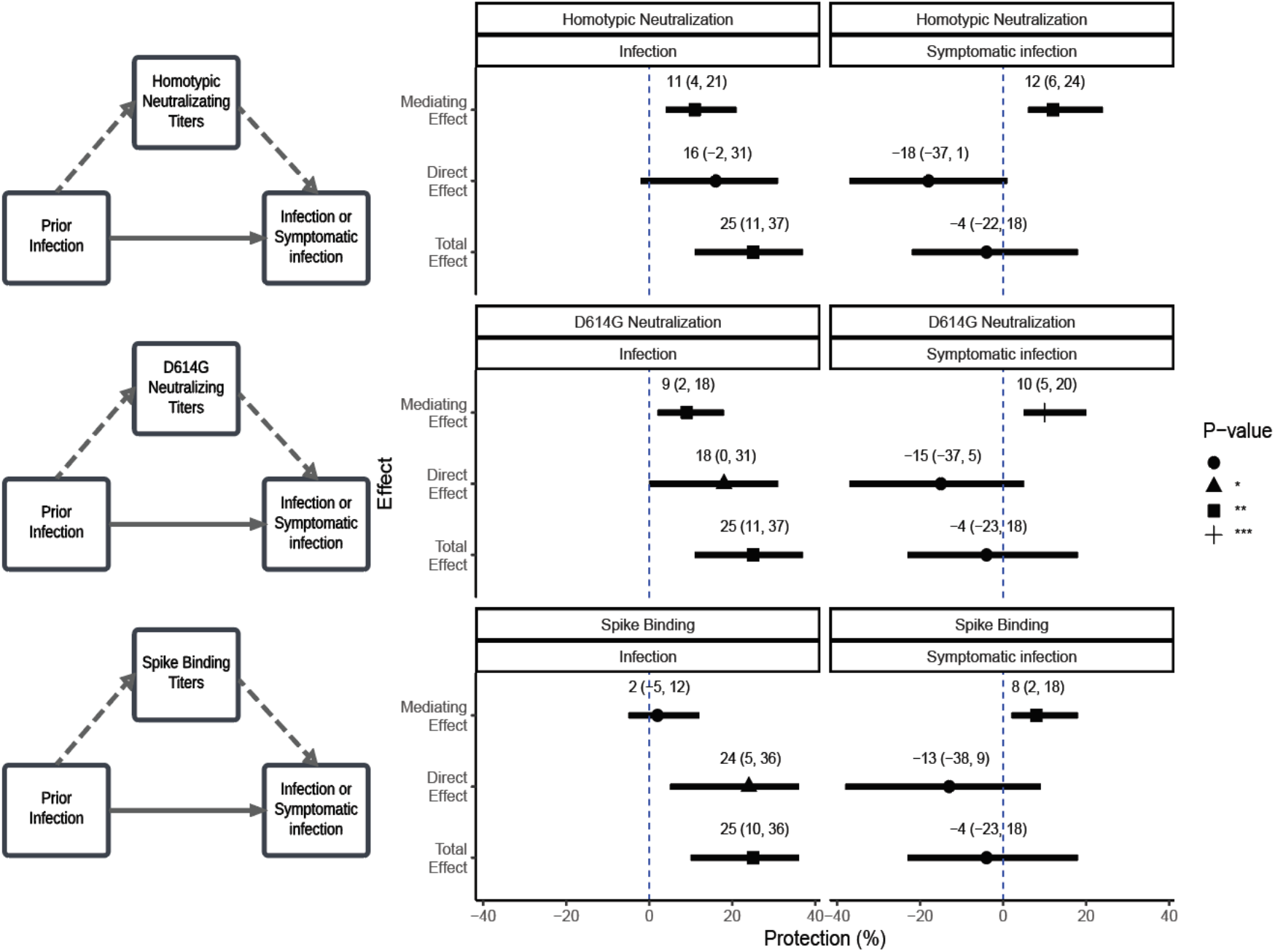
Mediation analysis of antibody effects on infection and symptomatic infection through prior infection. Each mediation analysis (across panels) were adjusted for age and prior vaccination or prior infection accordingly. This figure illustrates the total effect, average direct effect (solid lines), and average causal mediating effect (dashed lines) of antibody responses from any prior infection on protection against infection and symptomatic infection. Protection effects are expressed as percentages ([1-RR] x100). Dashed line indicates null effect. P-values are indicated for statistical significance, with ***, and * corresponding to P-values of <0.001, <0.01, and <0.05, respectively.

## Discussion

In this carefully monitored cohort of household participants in Managua, Nicaragua, we found that both homotypic Omicron-neutralizing antibody titers (e.g., anti-BA.1 titer for BA.1 wave participants) and ancestral SARS-CoV-2-neutralizing antibody titers were informative correlates of protection against symptomatic infection during Omicron BA.1 and BA.2 infection waves in 2022. Mediation analysis further revealed that neutralizing antibodies causally mediate protection from infection conferred by vaccination or prior infection. Our findings confirm that neutralizing antibody titers serve as a meaningful correlate of protection, measuring a causally protective factor, that may inform risk assessment.

Many prior studies have demonstrated, to varying degrees, that humoral immunity serves as a correlate of protection from SARS-CoV-2 infection or severe symptoms.^2-7,9,15,21,22^ The results presented here align with previous findings on ancestral SARS-CoV-2-neutralizing titers but also extend beyond prior work in key ways. First, the participants in this study reside in Managua, Nicaragua where SARS-CoV-2 had circulated extensively before the Omicron waves. The original SARS-CoV-2 strain started circulation in 2020, by 2021 Gamma and Delta predominated, and starting 2022 Omicron took over, first BA.1 and then BA.2 quickly replaced BA.1.^23^ By the time of this study’s sampling, people had been infected multiple times and subsequently vaccinated.^16^ The vaccines administered in this population included Sputnik and AstraZeneca for adults, and Abdala and Soberana for children.^24^

To the best of our knowledge, no prior studies on neutralizing antibody correlates of protection from COVID-19 have focused on participants in Nicaragua, or any other Central American countries. One prior study from South America , which included participants from Chile and Peru, analyzed ancestral-neutralizing titers in a phase 3 trial of AZD1222 (ChAdOx1 nCoV-19) vaccine.^25^ Our study provides critical insights into protective immunity in a region that has been underrepresented in global SARS-CoV-2 research. Here, we considered Nicaraguan participants with a variety of protective exposure histories (prior infections and various vaccines) and provided mediation analysis results separated by exposure.

Second, we utilized a household-based cohort study with an embedded transmission study design triggered by known virus exposures. Compared to clinical trials or standard observational cohort studies, exposures to infected individuals are expected to be more consistent across participants in household studies and more closely temporally linked to the measurement of antibody titers. For example, in Sun et al., 2024, samples were collected several weeks before high incidences of Omicron infections.^15^ Therefore, the results herein may be more reflective of similar viral exposure challenges across participants than in prior work, and correlates of protection from this study may be useful when considering protection levels at the time of exposure.

Third, we measured titers against the ancestral virus, to which participants were originally exposed earlier in the pandemic and/or by vaccination, as well as against contemporaneous Omicron subvariants BA.1 and BA.2. Relatively few correlates of protection studies have been published with such concurrent analyses of neutralizing antibodies directed against different prior and contemporaneous variants, and none have been household-based cohort with an embedded transmission study triggered by defined index case exposures around the time of titer measurement like this study.^15,26,27^ One household-based longitudinal cohort study of unvaccinated people in South Africa, without the nested triggering of sample collection that we deployed, measured anti-D614G and anti-BA.1 neutralizing titers months prior to a BA.1 infection wave. In that study, the authors found that BA.1-neutralizing titers mediated protection from infection elicited by prior infections, but also that the difference in neutralizing titer between D614G and BA.1 was itself no longer a correlate of protection, suggesting that homotypic-neutralizing antibody components of ancestral-neutralizing titers are the basis of the nAb correlate of protection.^15^ Our results showed that protective levels of D614G-neutralizing antibodies are likely higher than protective levels of homotypic-neutralizing antibodies, concordant with the results of Sun and colleagues, in both unvaccinated and vaccinated individuals. Intriguingly, our GLM-based correlate of protection analysis found that 4-fold increases in ancestral- and homotypic-neutralizing antibodies provided essentially equivalent increases in protection, despite higher average titers across the cohort directed against the ancestral strain. This result may also support the notion that during BA.1 and BA.2 waves, a fraction of cross-reactive ancestral-neutralizing antibodies also had activity against Omicron subvariants. Further, this result points to ancestral-neutralizing antibody titer being a useful correlate of protection even for a viral challenge with later Omicron BA.1 or BA.2 variants.

Our results are also consistent with literature showing that neutralizing antibodies causally mediate protection from infection by SARS-CoV-2 conferred by prior exposures. Here we further show that neutralizing antibodies mediate protection against infection, and more dramatically, protection against symptomatic infection, conferred by vaccination or prior infection. The cohort studied herein had exposure to global vaccines less frequently studied and most individuals had one or more infections before vaccination.

Finally, we show that serum antibodies, in particular neutralizing antibodies, are clearly informative correlates of protection from symptomatic infection. However, they were correlated with protection from asymptomatic infections only at higher titers, as detected by multiple prospective peri-exposure nucleic acid amplification tests paired with symptom questionnaires. This finding aligns with the pathophysiology of COVID-19 and the known limitations of serum antibodies, as mucosal immunity plays a key role in protection against initial infection.^28^ Thus, while serum antibody measurements are valuable for predicting protection against symptomatic COVID-19, they do not equally correlate with sterilizing immunity against overall SARS-CoV-2 infection.

This study has several limitations. First, there is potential ascertainment bias in our household-based cohort study design, as households with higher susceptibility to viral transmission and disease may have had a greater likelihood of being included due to the occurrence of an index case. Second, while we utilized neutralizing antibody assays to establish protective levels, variations in assay methodologies across different clinical laboratories may influence the exact protective thresholds reported, limiting cross-study comparisons. Third, our analysis to detect correlates of protection for BA.1 infection may be underpowered. Fourth, the assumption in our analyses that all household members experienced equivalent exposure to the virus may not fully capture variability in individual exposure risks within households. Finally, while our causal mediation analysis supports the role of neutralizing antibodies in mediating protection, the underlying assumptions of the mediation model cannot be fully verified. Therefore, causal interpretations of these results should be made cautiously.

In conclusion, in this household-based cohort with an embedded transmission study conducted in Managua, Nicaragua, we demonstrated that neutralizing antibody titers, particularly against future strains, are informative correlates of protection from infection and symptomatic SARS-CoV-2 infection. Mediation analyses provide evidence that neutralizing antibodies contribute to protection conferred by vaccination or prior infection, underscoring their role as a mechanistic correlate of immunity. Our findings emphasize the need for continuous surveillance of immune responses to evolving viral variants.

## Data Availability

Researchers interested in accessing the study data are encouraged to submit a formal request to A.G. or the Committee for the Protection of Human Subjects at the University of Michigan. To uphold ethical standards and ensure appropriate data use, each request will undergo a case-by-case review and approval process. Additionally, as the data include information collected in Nicaragua, access is subject to Nicaraguan data ownership regulations and may require approval from relevant Nicaraguan authorities.

## Notes

## Author contributions

J.V.Z.: conceptualization, data curation, formal analysis, methodology, software, validation, visualization, writing—original draft, writing—review and editing. I.A.M: investigation, formal analysis, methodology, writing—original draft, writing—review and editing. A.S.: data curation, methodology, writing—review and editing. H.E.M: data curation, methodology, writing—review and editing. M.P. and G.K: investigation, project administration, writing—review and editing. N.S.: investigation, writing—review and editing. Q.W.: investigation, writing—review and editing. L.L.: investigation, writing—review and editing. A.B.: investigation, methodology, supervision, project administration, writing—review and editing.

D.D.H: conceptualization, investigation, methodology, project administration, resources, supervision, writing—review and editing. A.G.: conceptualization, funding acquisition, investigation, methodology, project administration, resources, supervision, writing—review and editing.

## Acknowledgments

We thank the study personnel at the Centro de Salud Sócrates Flores Vivas, the Nicaraguan National Virology Laboratory, and the Sustainable Sciences Institute in Nicaragua. We are particularly grateful to the study participants and their families.

## Transparency declarations

A.G. has received institutional funding from Flu Lab and Open Philanthropy; personal honoraria from Hope College and the La Jolla Institute of Immunology; compensation for expert testimony from Berman and Simmons; and travel support from the Gates Foundation and the National Institutes of Health (NIH). A.G. has also served or serves in an advisory capacity to Janssen Pharmaceuticals and Sanofi Pasteur. D.D.H. co-founded TaiMed Biologics and RenBio, and he serves as a consultant for WuXi Biologics and Brii Biosciences and is a board director at Vicarious Surgical. All other authors declare no potential competing interests.

## References

1. Zohar T, Loos C, Fischinger S, et al. Compromised Humoral Functional Evolution Tracks with SARS-CoV-2 Mortality. Cell 2020; 183(6): 1508-19.e12.

2. Khoury DS, Cromer D, Reynaldi A, et al. Neutralizing antibody levels are highly predictive of immune protection from symptomatic SARS-CoV-2 infection. Nat Med 2021; 27(7): 1205–11.

3. Khoury DS, Schlub TE, Cromer D, et al. Correlates of Protection, Thresholds of Protection, and Immunobridging among Persons with SARS-CoV-2 Infection. Emerg Infect Dis 2023; 29(2): 381–8.

4. Goldblatt D, Alter G, Crotty S, Plotkin SA. Correlates of protection against SARS-CoV-2 infection and COVID-19 disease. Immunol Rev 2022; 310(1): 6–26.

5. Gilbert PB, Montefiori DC, McDermott AB, et al. Immune correlates analysis of the mRNA-1273 COVID-19 vaccine efficacy clinical trial. Science 2022; 375(6576): 43–50.

6. Feng S, Phillips DJ, White T, et al. Correlates of protection against symptomatic and asymptomatic SARS-CoV-2 infection. Nature Medicine 2021; 27(11): 2032–40.

7. Fong Y, Huang Y, Benkeser D, et al. Immune correlates analysis of the PREVENT-19 COVID-19 vaccine efficacy clinical trial. Nat Commun 2023; 14(1): 331.

8. Feng S, Phillips DJ, White T, et al. Correlates of protection against symptomatic and asymptomatic SARS-CoV-2 infection. Nat Med 2021; 27(11): 2032–40.

9. Fong Y, McDermott AB, Benkeser D, et al. Immune correlates analysis of the ENSEMBLE single Ad26.COV2.S dose vaccine efficacy clinical trial. Nat Microbiol 2022; 7(12): 1996–2010.

10. Perry J, Osman S, Wright J, et al. Does a humoral correlate of protection exist for SARS-CoV-2? A systematic review. PLoS One 2022; 17(4): e0266852.

11. Wang Q, Iketani S, Li Z, et al. Alarming antibody evasion properties of rising SARS-CoV-2 BQ and XBB subvariants. Cell 2023; 186(2): 279–86 e8.

12. Liu L, Iketani S, Guo Y, et al. Striking antibody evasion manifested by the Omicron variant of SARS-CoV-2. Nature 2022; 602(7898): 676–81.

13. Wang P, Nair MS, Liu L, et al. Antibody resistance of SARS-CoV-2 variants B.1.351 and B.1.1.7. Nature 2021; 593(7857): 130–5.

14. Cromer D, Reynaldi A, Mitchell A, et al. Predicting COVID-19 booster immunogenicity against future SARS-CoV-2 variants and the benefits of vaccine updates. Nat Commun 2024; 15(1): 8395.

15. Sun K, Bhiman JN, Tempia S, et al. SARS-CoV-2 correlates of protection from infection against variants of concern. Nat Med 2024; 30(10): 2805–12.

16. Maier HE, Balmaseda A, Saborio S, et al. Protection Associated with Previous SARS-CoV-2 Infection in Nicaragua. N Engl J Med 2022; 387(6): 568–70.

17. Chu DKW, Pan Y, Cheng SMS, et al. Molecular Diagnosis of a Novel Coronavirus (2019-nCoV) Causing an Outbreak of Pneumonia. Clin Chem 2020; 66(4): 549–55.

18. Maier HE, Plazaola M, Lopez R, et al. SARS-CoV-2 infection-induced immunity and the duration of viral shedding: Results from a Nicaraguan household cohort study. Influenza Other Respir Viruses 2023; 17(1): e13074.

19. Tingley D, Yamamoto T, Hirose K, Keele L, Imai K. mediation: R Package for Causal Mediation Analysis. Journal of Statistical Software 2014; 59(5).

20. Team RC. R: A Language and Environment for Statistical Computing. 2023.

21. Regev-Yochay G, Lustig Y, Joseph G, et al. Correlates of protection against COVID-19 infection and intensity of symptomatic disease in vaccinated individuals exposed to SARS-CoV-2 in households in Israel (ICoFS): a prospective cohort study. Lancet Microbe 2023; 4(5): e309–e18.

22. Woudenberg T, Pinaud L, Garcia L, et al. Estimated protection against COVID-19 based on predicted neutralisation titres from multiple antibody measurements in a longitudinal cohort, France, April 2020 to November 2021. Euro Surveill 2023; 28(25).

23. Aleman GV, Cerpas C, Juarez JG, et al. Tracking the genetic diversity of SARS-CoV-2 variants in Nicaragua throughout the COVID-19 Pandemic. bioRxiv 2024.

24. Zambrana JV, Saenz C, Maier HE, et al. Comparative Analysis of SARS-CoV-2 Antibody Responses across Global and Lesser-Studied Vaccines. Vaccines (Basel) 2024; 12(3).

25. Benkeser D, Fong Y, Janes HE, et al. Immune correlates analysis of a phase 3 trial of the AZD1222 (ChAdOx1 nCoV-19) vaccine. NPJ Vaccines 2023; 8(1): 36.

26. Atti A, Insalata F, Carr EJ, et al. Antibody correlates of protection against Delta infection after vaccination: A nested case-control within the UK-based SIREN study. J Infect 2023; 87(5): 420–7.

27. Hertz T, Levy S, Ostrovsky D, et al. Correlates of protection for booster doses of the SARS-CoV-2 vaccine BNT162b2. Nat Commun 2023; 14(1): 4575.

28. Havervall S, Marking U, Svensson J, et al. Anti-Spike Mucosal IgA Protection against SARS-CoV-2 Omicron Infection. N Engl J Med 2022; 387(14): 1333–6.

