## Supplementary Material for "Variant-specific antibody correlates of protection against SARS-CoV-2 Omicron symptomatic and overall infections"

### Supplementary methods

*Infection histories*

We employed positive serology screens, antibody titer measurements (including four-fold increases), and RT-PCR positivity to identify primary and secondary infections. To qualify as an infection, serological samples had to be from participants aged at least 6 months to avoid misclassification of maternal antibodies. First infections confirmed by RBD screening or spike titer were required to precede the individual's initial COVID-19 vaccination. Serologically distinct infections were defined as those occurring ≥6 months apart, and secondary infections detected via spike titer could not include samples collected around the time of vaccination. For RT-PCR-based detection, infections had to be ≥60 days apart, and sequential infections identified by RT-PCR and a four-fold increase in titer had to be >90 days apart. Any infections identified within a 60-day window were flagged for manual review. For this study, we categorized infection histories by any prior infection vs natural infection naïve. Only RT-PCR-confirmed infections were used to evaluate BA.1 or BA.2 infections.

*Laboratory methods*

**Cell lines.** Vero-E6 (CRL-1586) cells and HEK293T (CRL-3216) cells were obtained from ATCC and cultured at 37°C plus 5% $CO_2$ in Dulbecco's Modified Eagle Medium (DMEM, Sigma XXX) + 10% Fetal Bovine Serum (FBS, Sigma XXX) + 1% penicillin-streptomycin (Sigma XXX). Vero-E6 cells are derived from African green monkey kidneys. HEK293T cells are female human in origin.

**Pseudovirus production.** VSV-based SARS-CoV-2 pseudoviruses, in which the native VSV glycoprotein was replaced by SARS-CoV-2 spike and its variants, were produced as previously described. Plasmids containing the appropriate spike were transfected into HEK293T cells with PEI. 24 hours later, VSV-G pseudotyped ΔG-luciferase (G*ΔG-luciferase, Kerafast) was added to the cells, and then washed with culture medium three times before being cultured in fresh medium for another 24 hours. Anti-VSVG (I1) antibody was added to deplete non-pseudotyped viruses. Pseudoviruses were then harvested, centrifuged, aliquoted, and stored at -80°C.

**Pseudovirus titration.** All serum samples were heat inactivated at 56°C for 30 min before use. Pseudovirus particles bearing various SARS-CoV-2 spike proteins were inoculated onto Vero-E6 cells, starting with 50 µl per well in 96-well plates and then subjected to serial dilutions. Following a 16–18-hour incubation at 37°C, the activity of the virus-encoded firefly luciferase in cell lysates was measured using a SpectraMax luminometer (XXX). Each SARS-CoV-2 pseudovirus was titrated to standardize viral infectious dose before use in neutralization assays. The 50% tissue culture infectious dose ($TCID_{50}$) was calculated for each pseudovirus and used in all subsequent neutralization experiments.

**Pseudovirus serum neutralization assays.** Serially diluted (seven dilutions of) heat-inactivated sera were added in 96-well plates, starting at 1:100 dilution. Pseudoviruses were added and incubated at 37 °C for 1 hour. In each plate, wells containing only pseudoviruses were included as controls. Vero-E6 cells were then added at a density of $4 \times 10^4$ cells per well and incubate at 37 °C for an additional 16 hours. Cells were lysed and luminescence was determined by the Luciferase Assay System (Promega) and SoftMax Pro v.7.0.2 (Molecular Devices) according to the manufacturers' instructions. Data were analyzed in R v4.3.2, and ID50s were calculated using a 5-parameter log-logistic model with drda v2.0.3.

### Supplementary tables

**Table S1.** Univariate analysis by Infection, assay and Omicron wave.

| Omicron wave | Assay | Median difference* | statistic | P-value |
|---|---|---|---|---|
| BA.1 wave | BA.1 Neutralization, Log$_4$[ID$_{50}$] | 0.32 (-0.03, 0.75) | 8455.0 | 0.0878 |
| BA.2 wave | BA.1 Neutralization, Log$_4$[ID$_{50}$] | 0.83 (0.36, 1.42) | 1549.0 | 0.000427 |
| BA.1 wave | BA.2 Neutralization, Log$_4$[ID$_{50}$] | 0.18 (-0.11, 0.56) | 8140.0 | 0.252 |
| BA.2 wave | BA.2 Neutralization, Log$_4$[ID$_{50}$] | 0.87 (0.34, 1.47) | 1539.0 | 0.000567 |
| BA.1 wave | D614G Neutralization, Log$_4$[ID$_{50}$] | 0.21 (-0.14, 0.55) | 8157.5 | 0.240 |
| BA.2 wave | D614G Neutralization, Log$_4$[ID$_{50}$] | 0.69 (0.24, 1.16) | 1481.0 | 0.00266 |
| BA.1 wave | Spike Binding, Log$_4$[Titer] | 0 (-0.35, 0.34) | 7491.0 | 0.939 |
| BA.2 wave | Spike Binding, Log$_4$[Titer] | 0.49 (0.02, 1.01) | 1362.5 | 0.0357 |

*Estimate and 95% confidence interval

**Table S2.** Univariate analysis by symptomatic infections, assay and Omicron wave.

| Omicron wave | Assay | Median difference* | statistic | P-value |
|---|---|---|---|---|
| BA.1 wave | BA.1 Neutralization, Log$_4$[ID$_{50}$] | 0.6 (0.12, 1.05) | 8023.0 | 0.00937 |
| BA.2 wave | BA.1 Neutralization, Log$_4$[ID$_{50}$] | 1.02 (0.41, 1.79) | 1364.5 | 0.000559 |
| BA.1 wave | BA.2 Neutralization, Log$_4$[ID$_{50}$] | 0.38 (0, 0.81) | 7726.0 | 0.0417 |
| BA.2 wave | BA.2 Neutralization, Log$_4$[ID$_{50}$] | 0.98 (0.37, 1.64) | 1339.0 | 0.00119 |
| BA.1 wave | D614G Neutralization, Log$_4$[ID$_{50}$] | 0.3 (-0.07, 0.67) | 7505.5 | 0.106 |
| BA.2 wave | D614G Neutralization, Log$_4$[ID$_{50}$] | 0.79 (0.27, 1.32) | 1299.0 | 0.00357 |
| BA.1 wave | Spike Binding, Log$_4$[Titer] | 0.21 (-0.08, 0.64) | 7250 | 0.189 |
| BA.2 wave | Spike Binding, Log$_4$[Titer] | 0.67 (0.13, 1.17) | 1245.5 | 0.0133 |

*Estimate and 95% confidence interval

**Table S3 -** Multivariate analysis by infection status, Omicron wave and assay.

| Assay | Omicron wave | Variable | OR (95% CI) | Protection% (95% CI) | P-value |
|---|---|---|---|---|---|
| BA.1 Neutralization, $Log_4[ID_{50}]$ | BA.1 wave | Titer | 0.88 (0.73, 1.05) | 12 (-5, 27) | 0.160 |
| | | Prior Vaccination | 1.53 (0.85, 2.77) | -53 (-177, 15) | 0.160 |
| | | Prior infection | 0.31 (0.07, 1.03) | 69 (-3, 93) | 0.0804 |
| | | Age | 1.02 (1, 1.03) | -2 (-3, 0) | 0.0213 |
| | BA.2 wave | **Titer** | **0.55 (0.36, 0.79)** | **45 (21, 64)** | **0.00246** |
| | | Prior Vaccination | 0.98 (0.34, 2.91) | 2 (-191, 66) | 0.965 |
| | | Prior infection | 0.31 (0.01, 2.52) | 69 (-152, 99) | 0.326 |
| | | Age | 1 (0.98, 1.02) | 0 (-2, 2) | 0.870 |
| BA.2 Neutralization, $Log_4[ID_{50}]$ | BA.1 wave | Titer | 0.87 (0.72, 1.06) | 13 (-6, 28) | 0.169 |
| | | Prior Vaccination | 1.56 (0.85, 2.88) | -56 (-188, 15) | 0.149 |
| | | Prior infection | 0.31 (0.07, 1.06) | 69 (-6, 93) | 0.0869 |
| | | age | 1.02 (1, 1.03) | -2 (-3, 0) | 0.0163 |
| | BA.2 wave | **Titer** | **0.55 (0.36, 0.79)** | **45 (21, 64)** | **0.00281** |
| | | Prior Vaccination | 1 (0.35, 2.98) | 0 (-198, 65) | 0.998 |
| | | Prior infection | 0.33 (0.02, 2.98) | 67 (-198, 98) | 0.365 |
| | | Age | 1 (0.98, 1.02) | 0 (-2, 2) | 0.974 |
| D614G Neutralization, $Log_4[ID_{50}]$ | BA.1 wave | Titer | 0.87 (0.69, 1.09) | 13 (-9, 31) | 0.219 |
| | | Prior Vaccination | 1.56 (0.84, 2.9) | -56 (-190, 16) | 0.158 |
| | | Prior infection | 0.3 (0.07, 1.01) | 70 (-1, 93) | 0.0755 |
| | | Age | 1.02 (1, 1.03) | -2 (-3, 0) | 0.0151 |
| | BA.2 wave | **Titer** | **0.49 (0.29, 0.77)** | **51 (23, 71)** | **0.00383** |
| | | Prior Vaccination | 1.38 (0.46, 4.45) | -38 (-345, 54) | 0.574 |
| | | Prior infection | 0.24 (0.01, 1.83) | 76 (-83, 99) | 0.221 |
| | | Age | 1 (0.98, 1.02) | 0 (-2, 2) | 0.877 |
| Spike Binding, $Log_4[Titer]$ | BA.1 wave | Titer | 1.01 (0.85, 1.2) | -1 (-20, 15) | 0.938 |
| | | Prior Vaccination | 1.28 (0.71, 2.33) | -28 (-133, 29) | 0.412 |
| | | Prior infection | 0.24 (0.05, 0.83) | 76 (17, 95) | 0.0388 |
| | | Age | 1.02 (1, 1.03) | -2 (-3, 0) | 0.0112 |
| | BA.2 wave | Titer | 0.7 (0.45, 1.02) | 30 (-2, 55) | 0.0758 |
| | | Prior Vaccination | 1.04 (0.36, 3.13) | -4 (-213, 64) | 0.936 |
| | | Prior infection | 0.18 (0.01, 1.29) | 82 (-29, 99) | 0.132 |
| | | Age | 1 (0.98, 1.02) | 0 (-2, 2) | 0.971 |

**Table S4.** Multivariate analysis by symptomatic infection status, Omicron wave and assay.

| Assay | Omicron wave | Variable | OR (95% CI) | Protection (95% CI) | P-value |
|---|---|---|---|---|---|
| BA.1 Neutralization, $Log_4[ID_{50}]$ | BA.1 wave | **Titer** | **0.72 (0.58, 0.88)** | **28 (12, 42)** | **0.00164** |
| | | Prior Vaccination | 1.57 (0.82, 3.1) | -57 (-210, 18) | 0.181 |
| | | Prior infection | 3.9 (1.11, 18.6) | -290 (-1760, -11) | 0.0519 |
| | | age | 1.02 (1, 1.03) | -2 (-3, 0) | 0.0142 |
| | BA.2 wave | **Titer** | **0.53 (0.35, 0.76)** | **47 (24, 65)** | **0.000897** |
| | | Prior Vaccination | 2.11 (0.65, 8.29) | -111 (-729, 35) | 0.244 |
| | | Prior infection | 0.5 (0.05, 4) | 50 (-300, 95) | 0.518 |
| | | age | 0.99 (0.96, 1.01) | 1 (-1, 4) | 0.331 |
| BA.2 Neutralization, $Log_4[ID_{50}]$ | BA.1 wave | **Titer** | **0.72 (0.58, 0.89)** | **28 (11, 42)** | **0.00273** |
| | | Prior Vaccination | 1.65 (0.84, 3.31) | -65 (-231, 16) | 0.152 |
| | | Prior infection | 3.93 (1.11, 18.95) | -293 (-1795, -11) | 0.0522 |
| | | age | 1.02 (1.01, 1.03) | -2 (-3, -1) | 0.00786 |
| | BA.2 wave | **Titer** | **0.57 (0.38, 0.8)** | **43 (20, 62)** | **0.00220** |
| | | Prior Vaccination | 2.04 (0.63, 7.87) | -104 (-687, 37) | 0.261 |
| | | Prior infection | 0.55 (0.05, 5.11) | 45 (-411, 95) | 0.595 |
| | | age | 0.99 (0.97, 1.01) | 1 (-1, 3) | 0.455 |
| D614G Neutralization, $Log_4[ID_{50}]$ | BA.1 wave | **Titer** | **0.72 (0.56, 0.93)** | **28 (7, 44)** | **0.0123** |
| | | Prior Vaccination | 1.58 (0.8, 3.21) | -58 (-221, 20) | 0.191 |
| | | Prior infection | 3.4 (0.97, 16.22) | -240 (-1522, 3) | 0.0795 |
| | | age | 1.02 (1.01, 1.03) | -2 (-3, -1) | 0.00618 |
| | BA.2 wave | **Titer** | **0.43 (0.25, 0.71)** | 57 (29, 75) | 0.00157 |
| | | Prior Vaccination | 3.41 (0.94, 15.8) | -241 (-1480, 6) | 0.0840 |
| | | Prior infection | 0.39 (0.04, 3.05) | 61 (-205, 96) | 0.373 |
| | | age | 0.99 (0.97, 1.02) | 1 (-2, 3) | 0.532 |
| Spike Binding, $Log_4[Titer]$ | BA.1 wave | **Titer** | **0.82 (0.67, 0.99)** | **18 (1, 33)** | **0.0381** |
| | | Prior Vaccination | 1.37 (0.71, 2.69) | -37 (-169, 29) | 0.353 |
| | | Prior infection | 3.34 (0.92, 16.48) | -234 (-1548, 8) | 0.0922 |
| | | age | 1.02 (1.01, 1.04) | -2 (-4, -1) | 0.00164 |
| | BA.2 wave | **Titer** | **0.58 (0.36, 0.87)** | **42 (13, 64)** | **0.0131** |
| | | Prior Vaccination | 2.51 (0.75, 10.14) | -151 (-914, 25) | 0.158 |
| | | Prior infection | 0.27 (0.03, 1.93) | 73 (-93, 97) | 0.197 |
| | | age | 0.99 (0.97, 1.02) | 1 (-2, 3) | 0.587 |

**Table S5.** Protective thresholds – GAM model

| Assay | Omicron wave | 50% TS - Infection | 80% TS - Infection | 50% TS - Symptomatic infection | 80% TS - Symptomatic infection |
|---|---|---|---|---|---|
| BA.1 Neutralization, $ID_{50}$ | BA.1 Wave | - | - | 39 | 13107 |
| BA.2 Neutralization, $ID_{50}$ | | - | - | 53 | - |
| D614G Neutralization, $ID_{50}$ | | - | - | 222 | 78694 |
| Spike Binding, Titer | | - | - | 40 | - |
| BA.1 Neutralization, $ID_{50}$ | BA.2 Wave | 522 | 11809 | 186 | 3834 |
| BA.2 Neutralization, $ID_{50}$ | | 588 | 14685 | 181 | 5162 |
| D614G Neutralization, $ID_{50}$ | | 2276 | 33036 | 1145 | 11962 |
| Spike Binding, Titer | | 2581 | - | 1465 | 48560 |

TS: Threshold. Protective thresholds to achieve 50% or 80% protection.

**Table S6.** Protective thresholds – GLM model

| Assay | Omicron wave | 50% TS - Infection | 80% TS - Infection | 50% TS - Symptomatic infection | 80% TS - Symptomatic infection |
|---|---|---|---|---|---|
| BA.1 Neutralization, $ID_{50}$ | BA.1 Wave | - | - | 39 | 13107 |
| BA.2 Neutralization, $ID_{50}$ | | - | - | 53 | - |
| D614G Neutralization, $ID_{50}$ | | - | - | 222 | 78694 |
| Spike Binding, Titer | | - | - | 46 | - |
| BA.1 Neutralization, $ID_{50}$ | BA.2 Wave | 506 | 12540 | 186 | 3835 |
| BA.2 Neutralization, $ID_{50}$ | | 588 | 14685 | 181 | 5163 |
| D614G Neutralization, $ID_{50}$ | | 2276 | 33032 | 1188 | 11635 |
| Spike Binding, Titer | | 2581 | - | 1465 | 48560 |

TS: Threshold. Protective thresholds to achieve 50% or 80% protection.

**Table S7.** Mediation analyses

| Exposure | Mediator | Outcome | Term | RR (95%) | Protection (95% CI)* | P-value |
|---|---|---|---|---|---|---|
| Prior infection | Homotypic Neuts | Infection | Total Effect | 0.75 (0.63, 0.89) | 25 (37, 11) | 0.006 |
| | | | **ACME (average)** | **0.89 (0.79, 0.96)** | **11 (21, 4)** | **0.002** |
| | | | ADE (average) | 0.84 (0.69, 1.02) | 16 (31, -2) | 0.072 |
| | | Symptomatic Infection | Total Effect | 1.04 (0.82, 1.22) | -4 (18, -22) | 0.700 |
| | | | **ACME (average)** | **0.88 (0.76, 0.94)** | **12 (24, 6)** | **0.002** |
| | | | ADE (average) | 1.18 (0.99, 1.37) | -18 (1, -37) | 0.052 |
| Prior Vax | | Infection | Total Effect | 1.04 (0.93, 1.15) | -4 (7, -15) | 0.460 |
| | | | **ACME (average)** | **0.95 (0.91, 0.98)** | **5 (9, 2)** | **0.000** |
| | | | ADE (average) | 1.1 (0.98, 1.22) | -10 (2, -22) | 0.110 |
| | | Symptomatic Infection | Total Effect | 1.04 (0.94, 1.16) | -4 (6, -16) | 0.414 |
| | | | **ACME (average)** | **0.93 (0.89, 0.96)** | **7 (11, 4)** | **0.000** |
| | | | ADE (average) | 1.12 (1.02, 1.24) | -12 (-2, -24) | 0.016 |
| Prior infection | Neut Ancestral | Infection | Total Effect | 0.75 (0.63, 0.89) | 25 (37, 11) | 0.006 |
| | | | **ACME (average)** | **0.91 (0.82, 0.98)** | **9 (18, 2)** | **0.008** |
| | | | ADE (average) | 0.82 (0.69, 1) | 18 (31, 0) | 0.042 |
| | | Symptomatic Infection | Total Effect | 1.04 (0.82, 1.23) | -4 (18, -23) | 0.708 |
| | | | **ACME (average)** | **0.9 (0.8, 0.95)** | **10 (20, 5)** | **0.000** |
| | | | ADE (average) | 1.15 (0.95, 1.37) | -15 (5, -37) | 0.128 |
| Prior Vax | | Infection | Total Effect | 1.04 (0.93, 1.15) | -4 (7, -15) | 0.462 |
| | | | **ACME (average)** | **0.93 (0.88, 0.99)** | **7 (12, 1)** | **0.008** |
| | | | ADE (average) | 1.11 (0.99, 1.26) | -11 (1, -26) | 0.078 |
| | | Symptomatic Infection | Total Effect | 1.04 (0.94, 1.15) | -4 (6, -15) | 0.450 |
| | | | **ACME (average)** | **0.91 (0.85, 0.95)** | **9 (15, 5)** | **0.000** |
| | | | ADE (average) | 1.14 (1.04, 1.28) | -14 (-4, -28) | 0.002 |
| Prior infection | Spike Binding Ancestral | Infection | Total Effect | 0.75 (0.64, 0.9) | 25 (36, 10) | 0.002 |
| | | | ACME (average) | 0.98 (0.88, 1.05) | 2 (12, -5) | 0.574 |
| | | | ADE (average) | 0.76 (0.64, 0.95) | 24 (36, 5) | 0.018 |
| | | Symptomatic Infection | Total Effect | 1.04 (0.82, 1.23) | -4 (18, -23) | 0.656 |
| | | | **ACME (average)** | **0.92 (0.82, 0.98)** | **8 (18, 2)** | **0.006** |
| | | | ADE (average) | 1.13 (0.91, 1.38) | -13 (9, -38) | 0.206 |
| Prior Vax | | Infection | Total Effect | 1.04 (0.93, 1.15) | -4 (7, -15) | 0.514 |
| | | | ACME (average) | 0.99 (0.94, 1.04) | 1 (6, -4) | 0.574 |
| | | | ADE (average) | 1.05 (0.93, 1.17) | -5 (7, -17) | 0.412 |
| | | Symptomatic Infection | Total Effect | 1.03 (0.93, 1.14) | -3 (7, -14) | 0.558 |
| | | | **ACME (average)** | **0.94 (0.9, 0.98)** | **6 (10, 2)** | **0.006** |
| | | | ADE (average) | 1.09 (0.98, 1.21) | -9 (2, -21) | 0.114 |

*Protection calculated as (1-RR)x100.

### Supplementary figures

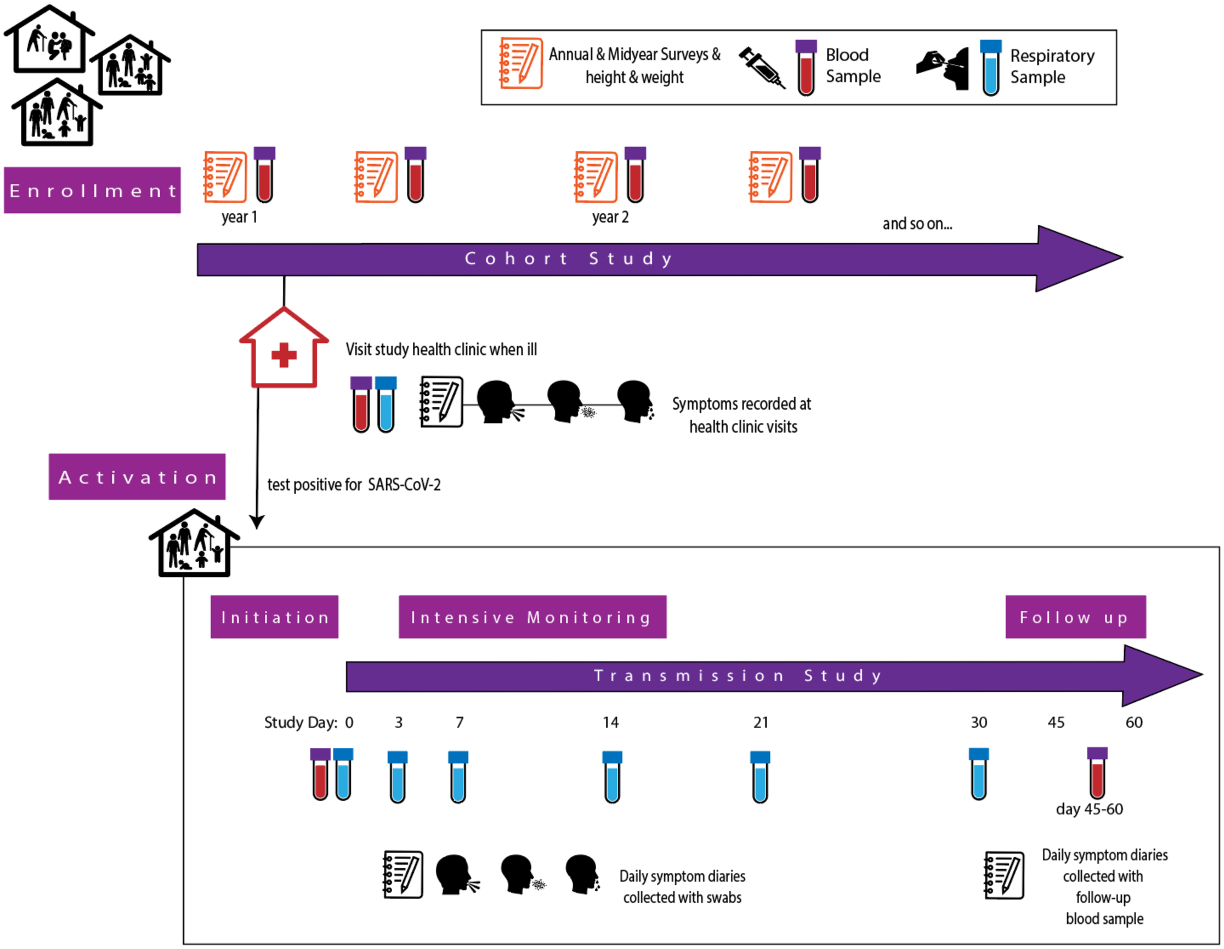


**Fig S1.** HICS Study design and nested transmission study design for SARS-CoV-2 activations.

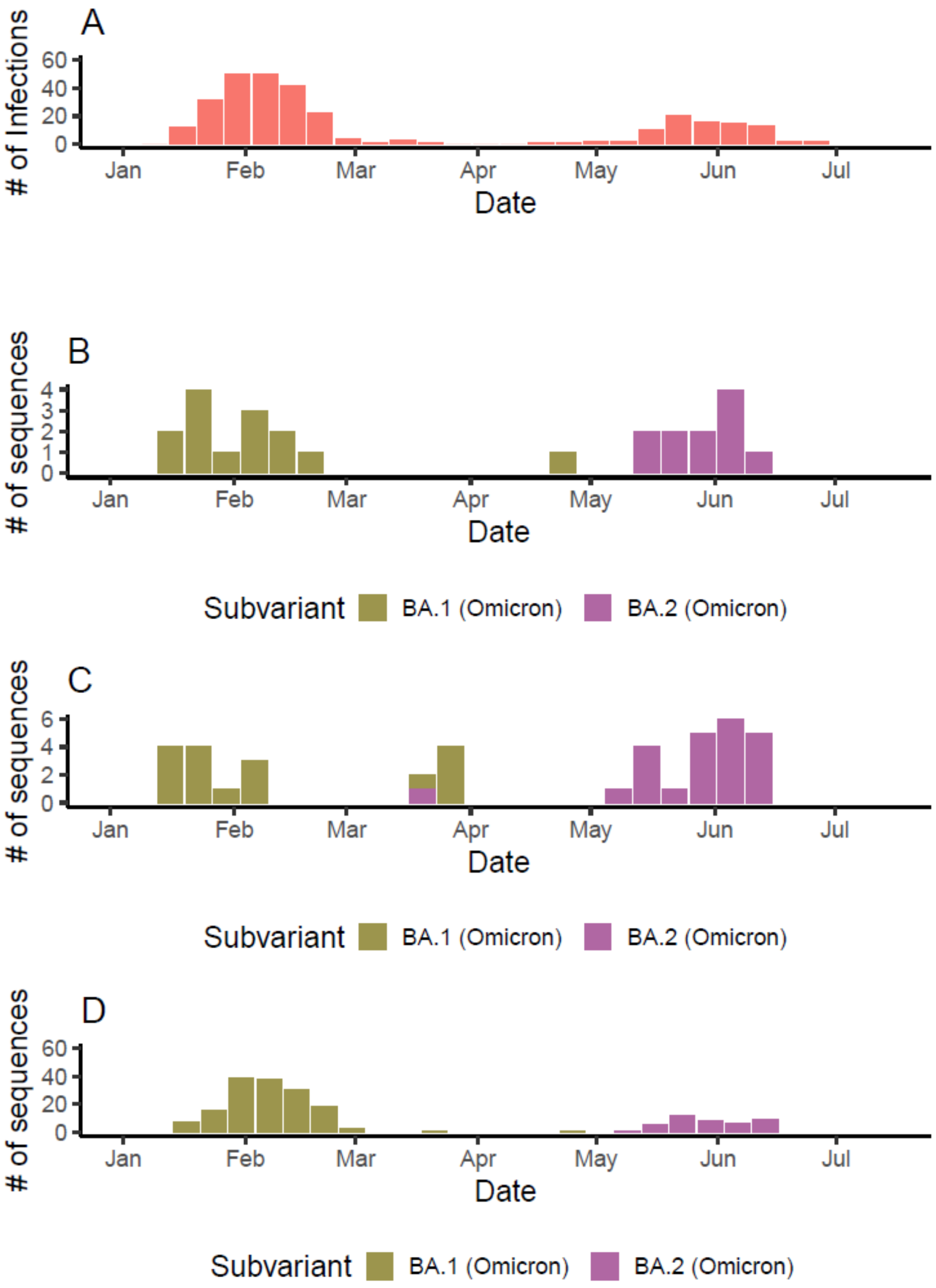


**Figure S2.** Infections in the HICS study, and sequencing data. (**A**) Total number of cases in the HICS study. (**B**) HICS individuals with sequencing data in the study period. (**C**) Sequenced patients from the Managua Department, extracted from: https://doi.org/10.1101/2024.06.03.596876. (**D**) Final sample with imputed sequencing data.

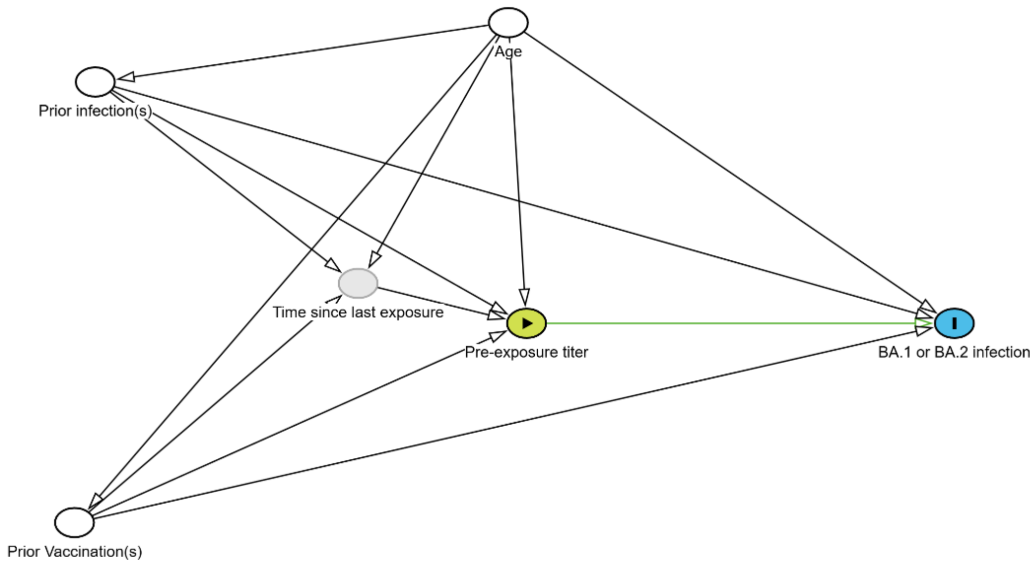


**Figure S3. Directed Acyclic Graph analysis.** This directed acyclic graph (DAG) illustrates the hypothesized relationships among key variables influencing susceptibility to BA.1 or BA.2 infection. Solid White nodes indicate variables adjusted for the analysis. The yellow node (Pre-exposure titer) is the principal variable of interest. The outcome is the blue node (BA.1 or BA.2 infection). The gray node is an unadjusted covariate. Solid black arrows depict putative causal pathways among variables adjusted for or considered potential confounders. The green arrow from Pre-exposure titer to BA.1 or BA.2 infection highlights the primary unbiased path of interest. Arrows flow from each variable to the factor or outcome it is hypothesized to influence.

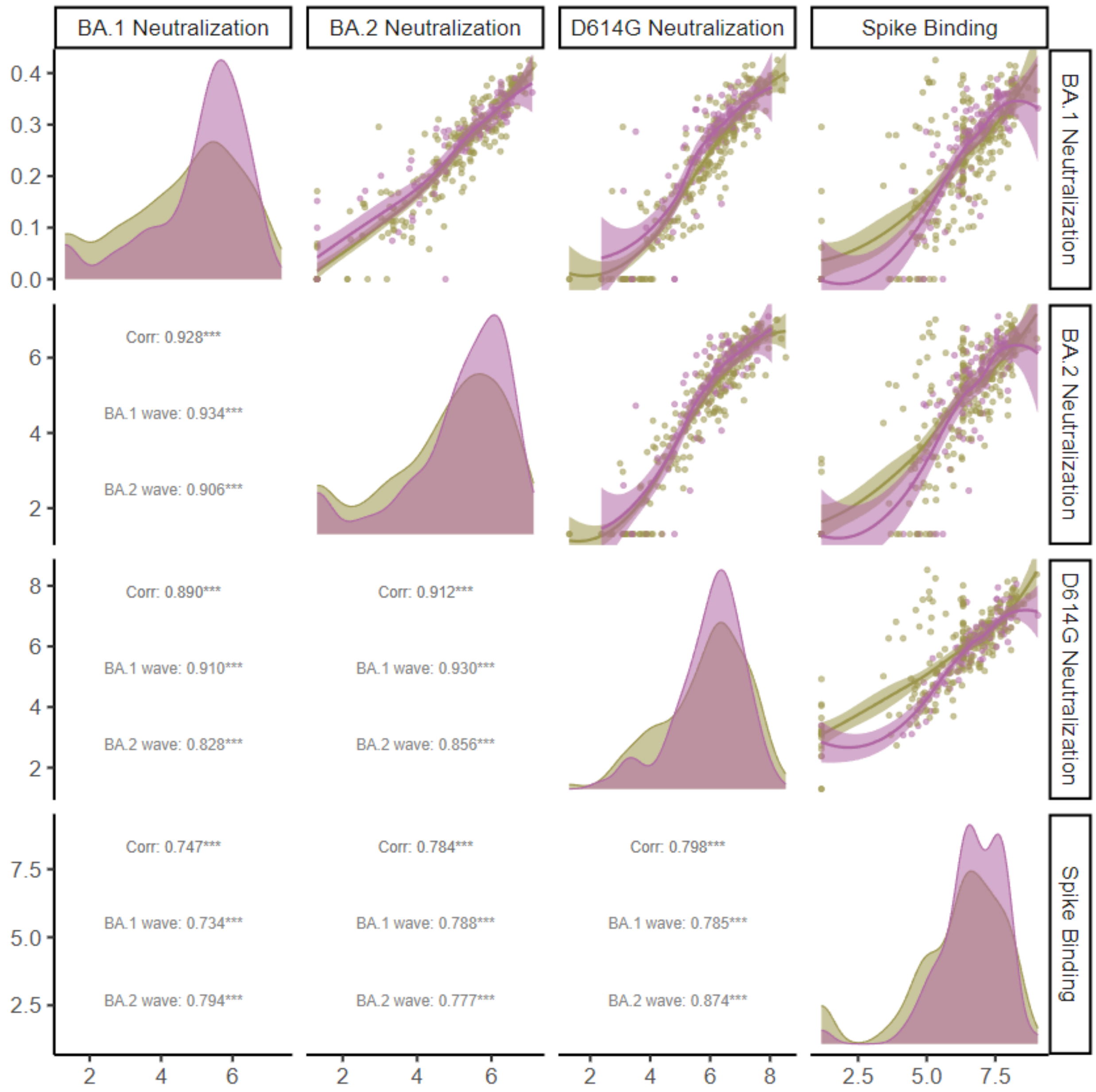


**Figure S4.** Correlations between assays. Pairwise correlations between BA.1 neutralization, BA.2 neutralization, D614G neutralization, and Spike-binding ELISA. Overall Pearson correlations are indicated as "Corr," while wave-specific correlations are labeled as "BA.1 wave" and "BA.2 wave." Correlation coefficients are displayed with levels of statistical significance: ***, **, and * corresponding to P-values of <0.001, <0.01, and <0.05, respectively. The diagonal histograms represent the titer distributions by omicron wave and assay. The distributions for the BA.1 wave are shown in golden-brown, while those for the BA.2 wave are depicted in purple. Each off-diagonal panel visualizes the relationship between two antibody responses, raw values and smooth LOESS correlations along with the 95% CI band colored in gray.

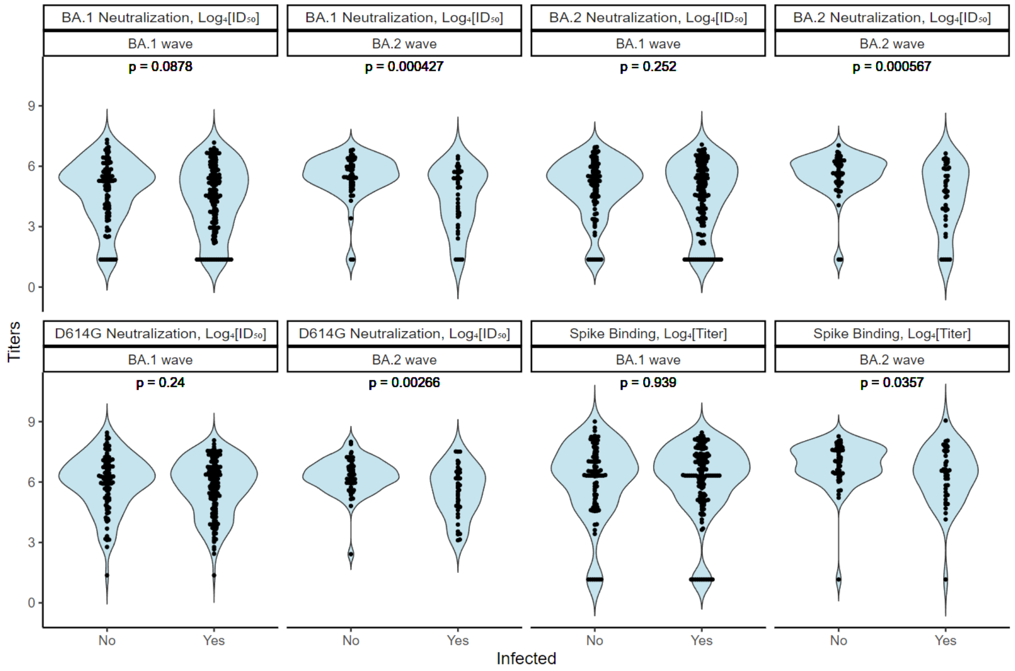


**Figure S5.** Univariate distributions by infection status. Univariate distributions of antibody responses by infection status during the BA.1 and BA.2 waves. P-values were calculated using the Wilcoxon rank-sum test.

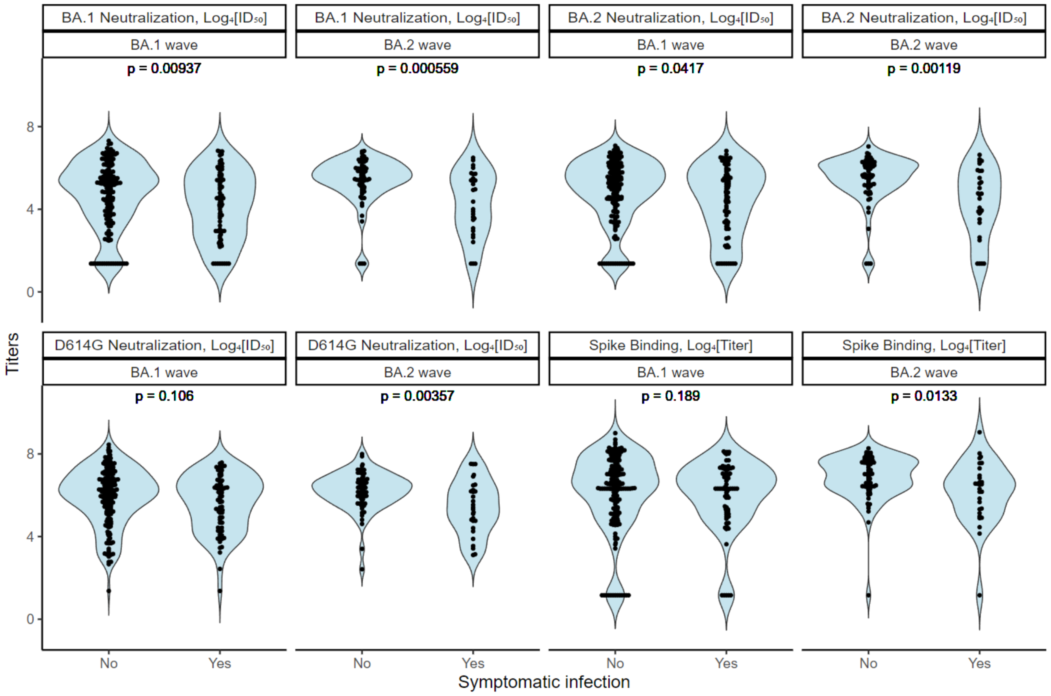


**Figure S6.** Univariate distributions by symptomatic infection status. Univariate distributions of antibody responses by symptomatic infection status during the BA.1 and BA.2 waves. P-values were calculated using the Wilcoxon rank-sum test.

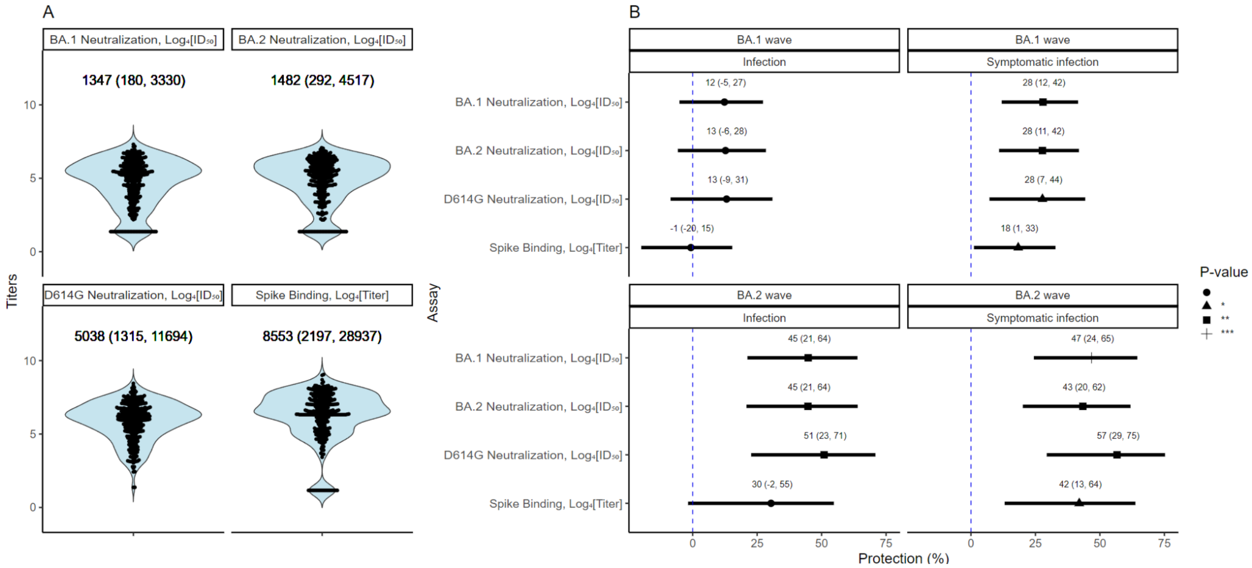


**Figure S7.** Univariate titer distributions and linear increase in neutralizing and binding titers as protective factors. (**A**) Univariate distributions by assay across Omicron waves. Untransformed median estimates and 25% and 75% percentiles are shown (**B**)This figure illustrates the effect of every four-fold rise in the likelihood of infection or symptomatic infection. Each estimate (per omicron wave, assay and outcome) is the result of single regression adjusting by age, any prior infection, and any prior vaccination. Odds ratio estimates were converted into protection estimates expressed as percentages ([1-OR] x 100). Dashed line indicates null effect. P-values are indicated for statistical significance, with ***, , and * corresponding to P-values of <0.001, <0.01, and <0.05, respectively.

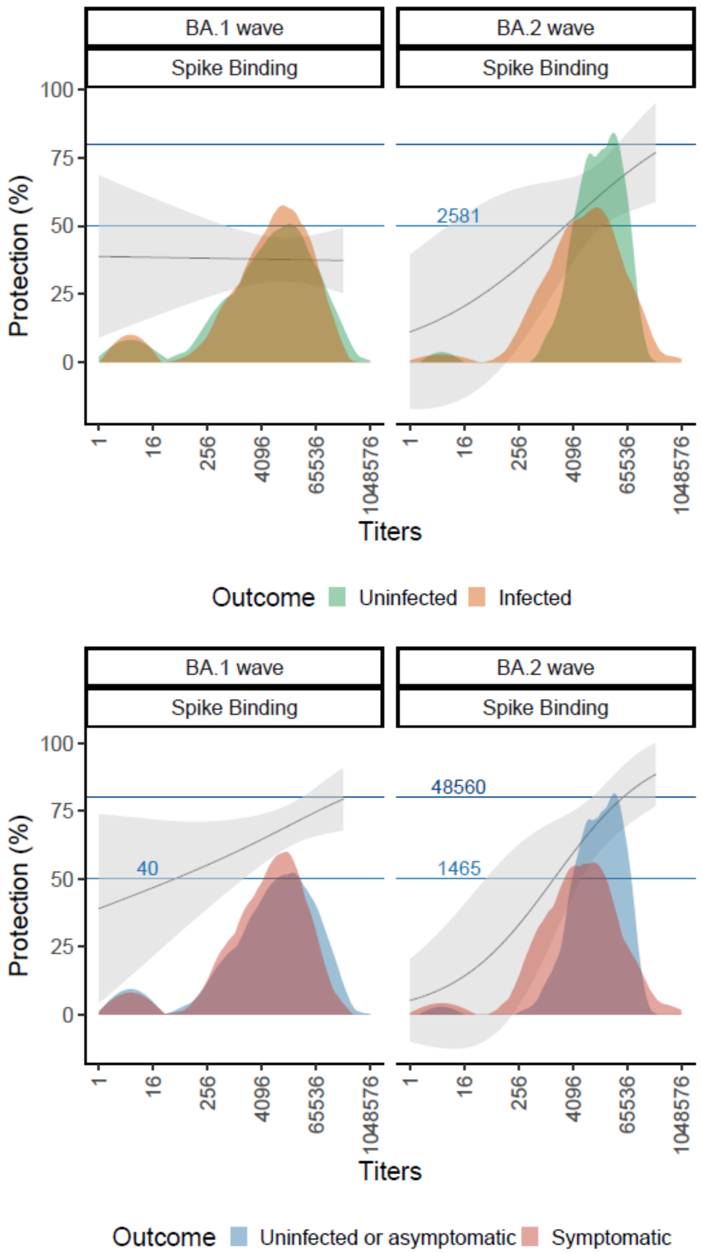


**Figure S8.** Protection curves for binding assays by Omicron wave and assay. This figure illustrates the relationship between binding titers and protection against BA.1 and BA.2 infection and symptomatic infection during their respective waves. The solid lines represent the predicted protection curves, with the shaded ribbon bands denoting the 95% confidence intervals (CIs) for these estimates, derived from vaccinated and prior-infected adults. The vertical dashed lines mark the points where the predicted protection curves cross the 50% and 80% protection thresholds, highlighting the titer levels required to achieve these levels of protection. Embedded within the figure are histograms showing the densities of neutralization titers, stratified by outcome (e.g., uninfected vs. infected or asymptomatic or uninfected vs. symptomatic).
